# Comorbid-phenome prediction and phenotype risk scores enhance gene discovery for generalized anxiety disorder and posttraumatic stress disorder

**DOI:** 10.1101/2021.07.13.21260369

**Authors:** Frank R Wendt, Gita A Pathak, Joseph D Deak, Flavio De Angelis, Dora Koller, Brenda Cabrera-Mendoza, Dannielle S Lebovitch, Daniel F Levey, Murray B Stein, Henry R Kranzler, Karestan C Koenen, Joel Gelernter, Laura M Huckins, Renato Polimanti

**Affiliations:** Department of Psychiatry, Yale School of Medicine, New Haven, CT, USA; VA CT Healthcare System, West Haven, CT, USA; Pamela Sklar Division of Psychiatric Genomics, Icahn School of Medicine at Mount Sinai, New York, NY 10029, USA; Department of Psychiatry, Icahn School of Medicine at Mount Sinai, New York, NY 10029, USA; Department of Genetics and Genomics, Icahn School of Medicine at Mount Sinai, New York, NY 10029, USA; Icahn Institute for Genomics and Multiscale Biology, Icahn School of Medicine at Mount Sinai, New York, NY 10029, USA; VA San Diego Healthcare System, Psychiatry Service, San Diego, CA, USA; Department of Psychiatry, University of California San Diego, La Jolla, CA, USA; Herbert Wertheim School of Public Health and Human Longevity Science, University of California San Diego, La Jolla, CA, USA; University of Pennsylvania Perelman School of Medicine, Philadelphia, PA, 19104, USA; Mental Illness Research, Education, and Clinical Center, Crescenz Veterans Affairs Medical Center, Philadelphia, PA, 19104, USA; Broad Institute of MIT and Harvard, Stanley Center for Psychiatry Research, Cambridge, MA, USA; Massachusettes General Hospital, Psychiatry and Neurodevelopmental Genetics Unit (PNGU), Boston, MA, USA; Harvard School of Public Health, Department of Epidemiology, Boston, MA, USA; Department of Genetics, Yale School of Medicine, New Haven, CT, USA; Department of Neuroscience, Yale School of Medicine, New Haven, CT, USA; Seaver Autism Center for Research and Treatment, Icahn School of Medicine at Mount Sinai, New York, NY 10029, USA; Mental Illness Research, Education and Clinical Center, James J. Peters Department of Veterans Affairs Medical Center, Bronx, NY 10468, USA

## Abstract

UK Biobank (UKB) is a key contributor in mental health genome-wide association studies (GWAS) but only ~31% of participants completed the Mental Health Questionnaire (“MHQ responders”). We predicted generalized anxiety disorder (GAD), posttraumatic stress disorder (PTSD), and major depression symptoms using elastic net regression in the ~69% of UKB participants lacking MHQ data (“MHQ non-responders”; N_Training_=50%; N_Test_=50%), maximizing the informative sample for these traits. MHQ responders were more likely to be female, from higher socioeconomic positions, and less anxious than non-responders. Genetic correlation of GAD and PTSD between MHQ responders and non-responders ranged from 0.636-1.08; both were predicted by polygenic scores generated from independent cohorts. In meta-analyses of GAD (*N*=489,579) and PTSD (*N*=497,803), we discovered many novel genomic risk loci (13 for GAD and 40 for PTSD). Transcriptomic analyses converged on altered regulation of prenatal dorsolateral prefrontal cortex in these disorders.

---

Psychiatric disorders are highly polygenic; thousands of risk loci across the genome contribute to their liability. Because of this polygenicity, extremely large sample sizes are required to detect the small individual effects associated with risk alleles.^1, 2, 3, 4, 5, 6^ Biobanks and consortia play a critical role in organizing, curating, and facilitating large genetic studies of mental health and psychopathology.^7, 8, 9, 10^ The UK Biobank (UKB) represents a resource of homogeneously ascertained participants with detailed information related to physical health, anthropometric measurements, and sociodemographic characteristics, etc. A primary limitation of UKB for studying mental health is the limited availability of participant responses to voluntary mental health questions and surveys. Among the approximately 502,000 UKB participants, only 31% completed the online Mental Health Questionnaire (herein termed “MHQ responders”).^11^ These missing data impose an upper limit on the UKB sample that is available for genetic studies using direct information. Indeed, many studies have had only modest success with risk locus discovery when studying psychopathologies in the subset of MHQ responders.^4, 12^

We hypothesized that carefully selected features ascertained in the entire UKB could permit genetic studies of MHQ phenotypes in the UKB participants who did not complete the survey (herein termed “MHQ non-responders”).^13^ We demonstrate the reliability of studying the collection of comorbid phenotypes, hereafter referred as a co-phenome,^13^ using several independent methods. Here we maximized the use of unrelated individuals from the UKB – more than doubling the available sample size relative to only MHQ responders – for genome-wide association studies (GWAS) of generalized anxiety disorder (GAD) and posttraumatic stress disorder (PTSD) symptoms. In meta-analyses adjusted for the effects of the major co-phenome correlate and an important transdiagnostic feature of internalizing psychopathologies, *neuroticism*, we identified multi-omic and cross-phenotype contributions of genes expressed in the prenatal brain. Using these novel GAD and PTSD data, we report putative cross-phenotype drug repurposing targets and identify drugs that may induce adverse effects that resemble anxiety symptoms. Our results provide one roadmap by which sample size and statistical power may be improved for gene discovery of incompletely ascertained traits in the UKB and other biobanks with limited mental health assessment.

## Results

A study overview is provided in Fig. 1.

**Fig. 1.**
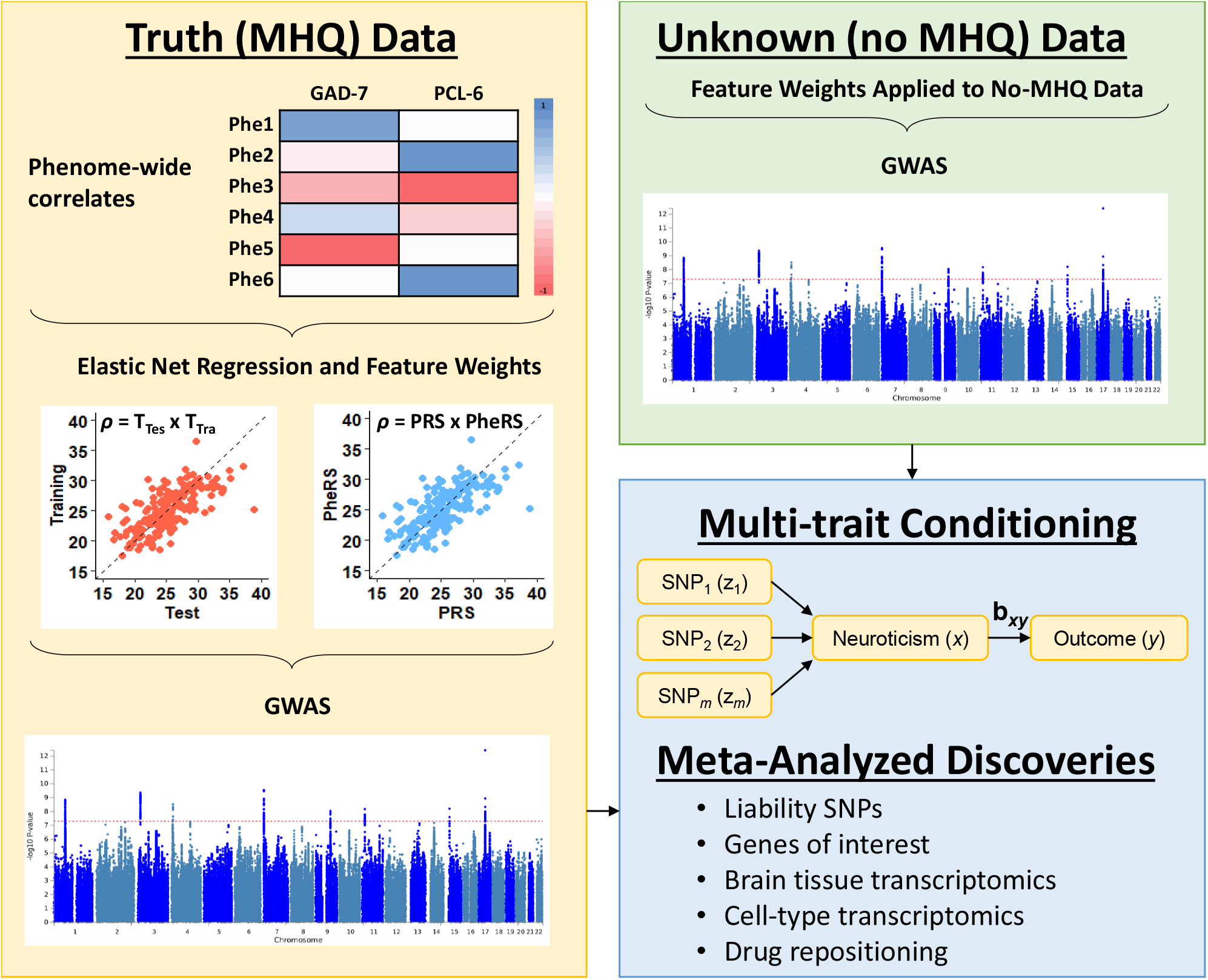
Study design for understanding the genetic architectures of internalizing co-phenomes. Features (i.e., comorbid phenotypes) were correlated with GAD-7, PCL-6, and PHQ-9. Outcomes were predicted using elastic net regression in two ways: (i) each quantitative outcome was predicted as the dependent variable in elastic net regression and (ii) elastic net regression weights were used to calculate a co-phenome risk score.

### Feature Selection and Elastic Net Regression

GAD-7 (GAD 7-item survey), PCL-6 (PTSD Checklist 6-item survey), and PHQ-9 (depression 9-item survey) quantitative scores were derived in MHQ responders according to previous studies^11^ (see Methods). After multiple testing correction for 772 phenotypes tested (see Methods; FDR<0.05), GAD-7, PCL-6, and PHQ-9 were correlated with 327, 363, and 373 phenotypes, respectively (Supplementary Table 1). We evaluated different combinations of training-test ratios and feature inclusion thresholds (defined as Spearman’s rho (*ρ*) relative to GAD-7, PCL-6, and PHQ-9; Supplementary Table 2).^14^ Among MHQ responders, correlation between observed and predicted GAD-7, PCL-6, and PHQ-9 across elastic net training and test ratios were consistent at all parameter tuning combinations. We predicted each outcome in MHQ non-responders using the elastic net regression parameters with the lowest root mean square error and highest magnitude of Spearman’s *ρ* between predicted and true outcomes. Using *ρ*>0.20 as a feature inclusion threshold, we predicted GAD-7 with 19 phenotypes (observed *versus* predicted among test data *ρ*=0.33, *P*<2×10^−16^), PCL-6 with 15 phenotypes (observed *versus* predicted among test data *ρ*=0.21, *P*<2×10^−16^), and PHQ-9 with 17 (observed *versus* predicted among test data *ρ*=0.33, *P*<2×10^−16^) phenotypes.

The correlated trait “*neuroticism score*” (Field ID 20127) was the feature that was most strongly associated with, and a major predictor of, internalizing symptoms in elastic net regression (“*neuroticism score*” versus GAD-7 *ρ*=0.482, *P*<4.13×10^−307^, elastic net *β*=0.286; “*neuroticism score*” versus PCL-6 *ρ*=0.378, *P*<4.13×10^−307^, elastic net *β*=0.103; “*neuroticism score*” versus PHQ-9 *ρ*=0.41, P<4.13×10^−307^, elastic net *β*=0.03). The remaining predictors of internalizing symptoms capture relevant relationships with these traits, including features such as “*tenseness*” (Field ID 1990), “*frequency of tiredness in the last two weeks*” (Field ID 2080), and “*overall health rating*” (Field ID 2178). Supplementary Table 3 shows all predictors and their elastic net regression weights in the UKB.

### Characteristics of MHQ responders and non-responders

Co-phenome risk scores (PheRS) are a weighted sum of the co-phenome questionnaire responses based on weights derived from elastic net regression described above. PheRS were more strongly correlated with predicted internalizing outcomes (MHQ non-responder data) than the same outcome directly ascertained in MHQ responders likely due to the independence of MHQ responder data relative to the lack of independence of these variables in MHQ non-responders (Supplementary Table 4). All predicted quantitative outcomes were greater in magnitude among the MHQ non-responders suggesting more severe symptoms compared to MHQ responders. While the difference was minor for PCL-6 (MHQ responder mean=6.69, s.d.=3.65; non-responder mean=6.82, s.d.=1.80; Cohen’s *d*=-0.048, *P*=9.96×10^−27^) and PHQ-9 (MHQ responder mean=11.73, s.d.=3.67; non-responder mean=12.02, s.d.=2.32; Cohen’s *d*=0.096, *P*=2.21×10^−144^), it was more pronounced for GAD-7 scores (MHQ responder mean=8.97, s.d.=3.09; non-responder mean=12.36, s.d.=4.78; Cohen’s *d*=-0.749, *P*=1×10^−322^).

In single-variable generalized linear models, “*neuroticism score*” (a summary score of neuroticism based on 12 neurotic behaviors; *β*=-0.080, s.e.=0.001, *P*<2×10^−16^), “*average household income before tax*” (*β*=0.239, s.e.=0.003, *P*<2×10^−16^), PCL-6 score (*β*=-0.052, s.e.=0.002, *P*<2×10^−16^), GAD-7 score (*β*=-0.202, s.e.=0.001, *P*<2×10^−16^), PHQ-9 score (*β*=-0.031, s.e.=0.002, *P*<2×10^−16^), and others were all associated with whether UKB participants responded to the MHQ (Supplementary Table 5). In a multivariable analysis of MHQ participation that also accounted for birthplace (north and east coordinates), age, and sex (full model *R*^*2*^=0.300, *P*<2.2×10^−16^), all variables remained significant (Supplementary Table 5). However, the effect of “*neuroticism score*” showed a significant opposite effect direction when accounting for the effects of the other variables included in the multivariable model (single-variable *β*=-0.080, s.e.=0.001, *P*<2×10^−16^; multivariable *β*=0.376, s.e.=0.003, *P*<2×10^−16^). This effect was due to the interplay of “*neuroticism score*” with GAD-7 symptoms (multivariable interaction term *neuroticism score*×GAD-7 *β*=-0.086, s.e.=0.001, *P*<2×10^−16^), but it was independent of PCL-6 and PHQ-9 symptoms. Upon excluding GAD-7 from the multivariable model, “*neuroticism score*” was a negative predictor of MHQ participation (*β*=-0.094, s.e.=0.002, *P*<2×10^−16^). Conversely, removing PCL-6 or PHQ-9 while keeping GAD-7 in the multivariable model, did not produce the same change (*β*=0.381, s.e.=0.003, *P*<2×10^−16^ and *β*=0.335, s.e.=0.003, *P*<2×10^−16^). Based on these observations, UKB participants with the highest “*neuroticism scores*” (i.e., 12; mean MHQ participation probability=97.7%, s.d.=0.151) were 6.04 times more likely to contribute to the MHQ than those with the lowest “*neuroticism score*” (i.e., 0; MHQ participation probability=16.2%, s.d.=0.872, *P*_diff_=1.03×10^−203^; Fig. 2). This effect appeared strongest among participants with medium (GAD-7=14) and low (GAD-7=7) GAD scores but was attenuated in the high GAD group (GAD-7=21). All samplings of GAD-7 and neuroticism score combinations are provided in Supplementary Table 5. Given the major contribution of “*neuroticism score*” in the elastic net prediction of GAD-7 (*ρ*_MHQ-responders_ =0.482, *P*<4.13×10^−307^; *ρ*_MHQ-non-responders_=0.984, *P*<4.13×10^−307^), this result may highlight residual participation bias transcending GAD, PTSD, and depression psychopathologies. We detected only modest evidence of multicollinearity in the model: *neuroticism score* variance inflation factor (VIF)=2.57, GAD-7 VIF=2.46, PCL-6 VIF=1.94, and PHQ VIF=2.27. A VIF > 10 is generally considered an indicator of high correlation and high multicollinearity.^15^ Because the correlation between GAD-7 and *neuroticism score* is modest (*ρ*=0.644), and the VIF for GAD-7 (2.46) and *neuroticism score* are relatively low, we can exclude multicollinearity is affecting our model of MHQ participation (Supplementary Table 5 and **Supplementary Results**).

**Fig 2.**
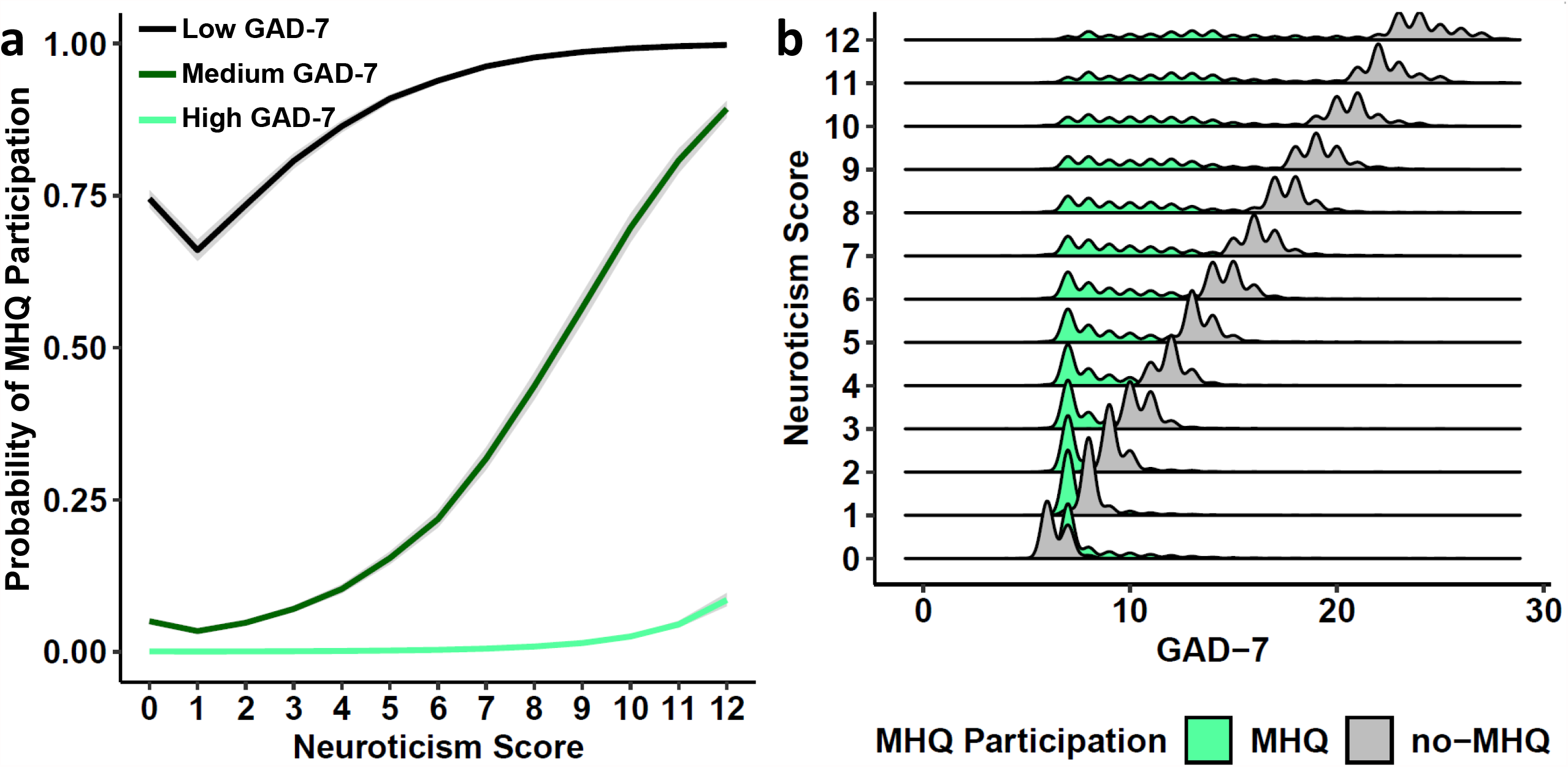
Probability of MHQ survey participation. (a) Probability of participating in the MHQ survey based on neuroticism score and three samplings of GAD-7 (low = 7, medium = 14, high = 21). In (b), the relationship between GAD-7 and neuroticism scores. All data describing these effects are provided in Supplementary Table 5.

### SNP-based Heritability

We describe below a series of tests to verify that elastic net-predicted GAD, PTSD, and depression outcomes and PheRS capture the same genetic liability as true observations of each outcome.

We performed three GWAS for each trait: quantitative score, PheRS, and binary case-control status derived from the quantitative scores. All GWAS included age, sex, age×sex, and ten within-ancestry principal components as covariates. While elastic-net prediction accuracies were relatively low, genetic analyses of predicted values captured similar information to that of direct-report internalizing phenotype scores. The SNP-based heritability (*h*^*2*^) for each GWAS was significantly different from zero (Supplementary Table 6). Due to the strong contribution of “*neuroticism score*” to elastic net predictions, the detected difference between MHQ responder and non-responder *neuroticism scores*, and the well-documented heritable and polygenic architecture of *neuroticism*,^16^ we analyze GWAS only after subjecting their effect sizes to multi-trait conditioning with a GWAS of *neuroticism*.^16, 17, 18^ This approach facilitated an understanding of how much polygenicity and GWAS biological annotation findings could be attributed to the relatively large heritable component of neuroticism versus that of the PheRS and quantitative traits. After conditioning, all GWAS still had *h*^*2*^ estimates that differed significantly from zero (GAD phenotype range: MHQ-responder GAD-7-PheRS [*h*^*2*^=0.81%, s.e.=0.40, *P*=0.043] to MHQ non-responder GAD-7-PheRS [*h*^*2*^=3.52%, s.e.=0.32, *P*=3.82×10^−28^]; PTSD phenotype range: MHQ responder PTSD [*h*^*2*^=1.88%, s.e.=0.21, *P*=3.55×10^−19^] to MHQ responder PCL-6 [*h*^*2*^=5.57%, s.e.=0.46, *P*=9.35×10^−35^]; depression phenotype range: MHQ non-responder current depression [*h*^*2*^=1.61%, s.e.=0.10, *P*=2.55×10^−58^] to MHQ non-responder PHQ-9 [*h*^*2*^=5.89%, s.e.=0.39, *P*=1.62×10^−51^]). Supplementary Table 6 shows *h*^*2*^ estimates for all traits before and after multi-trait conditioning with the GWAS of *neuroticism*. Unless otherwise noted, we focus all analyses on the internalizing traits that were subjected to multi-trait conditioning with *neuroticism*. However, the Supplementary Material corresponding to each analysis presents results prior to multi-trait conditioning so that comparisons and contrasts may be drawn. There were several *h*^*2*^ estimates that differed significantly between the MHQ responder and non-responder GWAS, but there was no evidence of a systematic over- or under-estimation of SNP-*h*^*2*^ in either cohort (Fig. 3 and Supplementary Table 6).

**Fig. 3.**
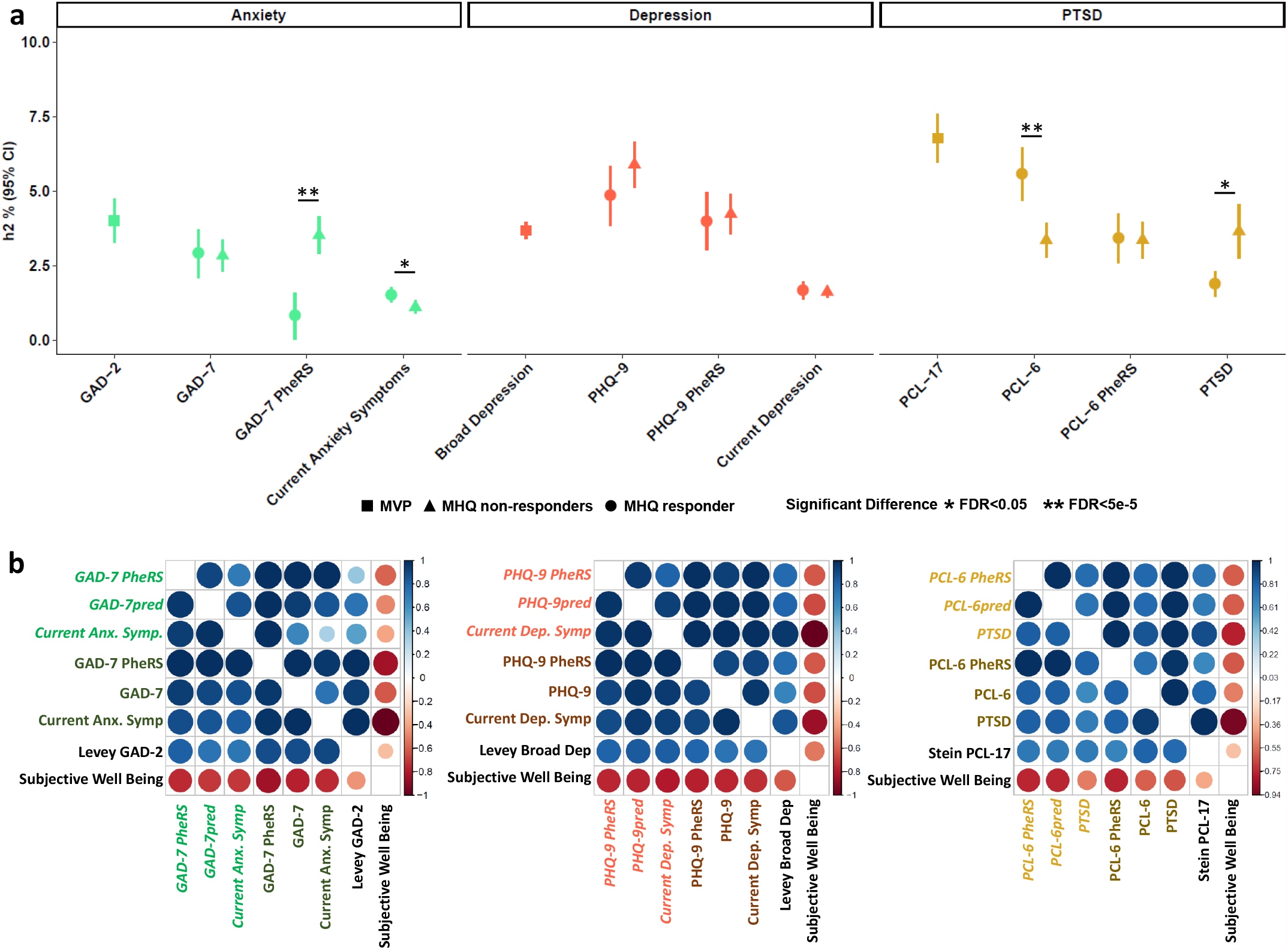
Verifying the concordant genetic architectures of true and predicted internalizing outcomes. **a**, SNP-heritability (*h*^*2*^) of each internalizing outcome and the current largest unrelated sampling of a corresponding phenotype (GAD-2, PCL-17, and broad depression) after multi-trait conditioning with neuroticism. Each data point is the trait h2 point estimate and error bars represent the 95% confidence interval (CI) associated with each estimate. **b**, Genetic correlation (*r*_*g*_) within and between internalizing outcomes derived from the Mental Health Questionnaire (MHQ responders) and those predicted in the MHQ non-responders of the UKB before (bottom left triangle) and after (top right triangle) multi-trait conditioning with neuroticism. Pale text indicates a phenotype from the MHQ non-responders and dark text indicates a phenotype from the MHQ responders. Each *r*_*g*_ heatmap contains a positive control with positive *r*_*g*_ (largest Million Veteran Program (MVP) corresponding phenotype) and positive control with negative *r*_*g*_ (subjective well-being) phenotype. All *r*_*g*_s survive multiple testing correction (*FDR*<0.05).

### Genetic Overlap between MHQ-responder and MHQ non-responder Traits

Genetic correlations (*r*_*g*_) between GWAS performed in MHQ responders and non-responders were statistically significant and relatively high when comparing the same trait: MHQ responder *versus* non-responder GAD-7-PheRS *r*_*g*_=1.55, s.e.=0.406, *P*=1.0×10^−4^; PCL-6-PheRS *r*_*g*_=1.19, s.e.=0.097, *P*=2.05×10^−34^; PHQ-9-PheRS *r*_*g*_=1.15, s.e.=0.084, *P*=3.52×10^−42^. The *r*_*g*_estimates among all other trait combinations are provided in Supplementary Table S7. Each UKB phenotype (assessed in MHQ responders and non-responders) was genetically correlated with the corresponding Million Veteran Program (MVP) phenotype Supplementary Table 7 and after multiple testing correction for the number of *r*_*gs*_ tested there were no differences in *r*_*g*_ with MVP phenotype between MHQ responders and non-responders. All internalizing outcomes were negatively genetically correlated with subjective well-being (*FDR*<0.05, Supplementary Table 7).

We next evaluated how genetic effects detected in the MVP cohort predicted internalizing outcomes in MHQ responders and non-responders (Fig. 3 and Supplementary Fig. 2). The MHQ non-responders generally had greater PRS z-scores and *R*^*2*^ relative to the responders (Supplementary Table 8). The PCL-6 phenotype variance explained was notably greater in the MHQ non-responders (e.g., MHQ responders minimum *P*-value=2.93×10^−21^, variance explained (*R*^*2*^)=0.063% and MHQ non-responders *P*-value=2.86×10^−40^, *R*^*2*^=0.092%, *P*_*T*_ =1×10^−5^ at *P*_*T*_=1×10^−5^) likely reflecting greater statistical power due to the PTSD informativeness of the MHQ non-responder sample. Regression coefficients for GAD and PTSD PheRS and predicted case-control status presented similar power improvements in the no-MHQ cohort (Supplementary Table 8). Due to complete sample overlap between UKB depression data and the MVP broad depression GWAS,^3^ PRS were not performed for predicted PHQ-9.

### Gene Discovery through Meta-Analysis

High genetic correlation and strong polygenic prediction between MHQ responder and non-responder GWAS support the reliability of the genetic information derived from MHQ non-responders. Accordingly, we combined the predicted data (MHQ non-responder) with the direct-report information (MHQ responder) available for GAD and PTSD. First, we meta-analyzed the UKB MHQ responders and non-responders to describe how using the entire UKB enhances gene discovery. Second, we meta-analyzed the two UKB cohorts with other available datasets. Here, we present data from the meta-analysis of UKB MHQ responders, non-responders, and MVP subjected to multi-trait conditioning with *neuroticism*. Results from meta-analyses not subjected to multi-trait conditioning with *neuroticism* are provided in **Supplementary Material** and should be interpreted with the expectation that much of the detected signal may be attributable to genetic liability to *neuroticism*. Results of meta-analyzed depression (UKB only) offered no increase in sample size or *h*^*2*^ relative to MVP broad depression,^3^ and therefore was omitted from *in silico* analyses.

Quantitative traits and PheRS revealed slightly different risk loci for each outcome (Supplemental Table 9 and Supplementary Fig. 1) but these differences may reflect random sampling noise as there was no statistical difference in the genome-wide significant variants effects (Supplementary Table 10). Applying a genome-wide multiple testing correction (*P*<5×10^−8^), we discovered (i) 10 and 12 risk loci for GAD when meta-analyzing with GAD-7 and GAD-7 PheRS, respectively and (ii) 32 and 26 risk loci for PTSD when meta-analyzing with PCL-6 and PCL-6 PheRS, respectively. We defined a credible set of likely causal loci: 70% of GAD-7 loci, 50% of GAD-7 PheRS loci, 46.8% of PCL-6 loci, and 23.1% of PCL-6 PheRS loci (Supplementary Table 11). Some loci that positionally mapped (2-kb windows; 1-kb window on both sides to capture cis-regulatory effects)^19^ to genic regions have prior evidence of involvement in GAD (*e*.*g*., *PHF2*-rs12376738 and resistance to depression- and anxiety-like symptoms^20^ and memory consolidation^21^), PTSD (*e*.*g*., *IL2*-rs45510091 and low dose cytokine treatments to reverse anxious symptoms), or related symptoms.^22^

After study-wide multiple testing correction (*P*_*adj*_<1.25×10^−8^ = 5×10^−8^/2 traits/2 definitions of each trait), 7 and 6 loci were associated with GAD based on meta-analyses with GAD-7 and GAD-7 PheRS, respectively and 22 and 19 loci were associated with PTSD based on meta-analyses with PCL-6 and PCL-6 PheRS, respectively (Fig. 4 and Supplementary Tables 9 and 10). Six genes were positionally mapped to genomic risk loci detected in GAD and PTSD GWAS: *ADAD1-IL2*-*IL21*-*KIAA1109* gene cluster, *CRHR1*-*MAPT*-*NSF*-*PLEKHM1*-*WNT3* gene cluster, *FAM120*-*FAM120AOS*-*PHF2* gene cluster, *MAD1L1, SOX6*, and *TMEM106B*.

**Fig. 4.**
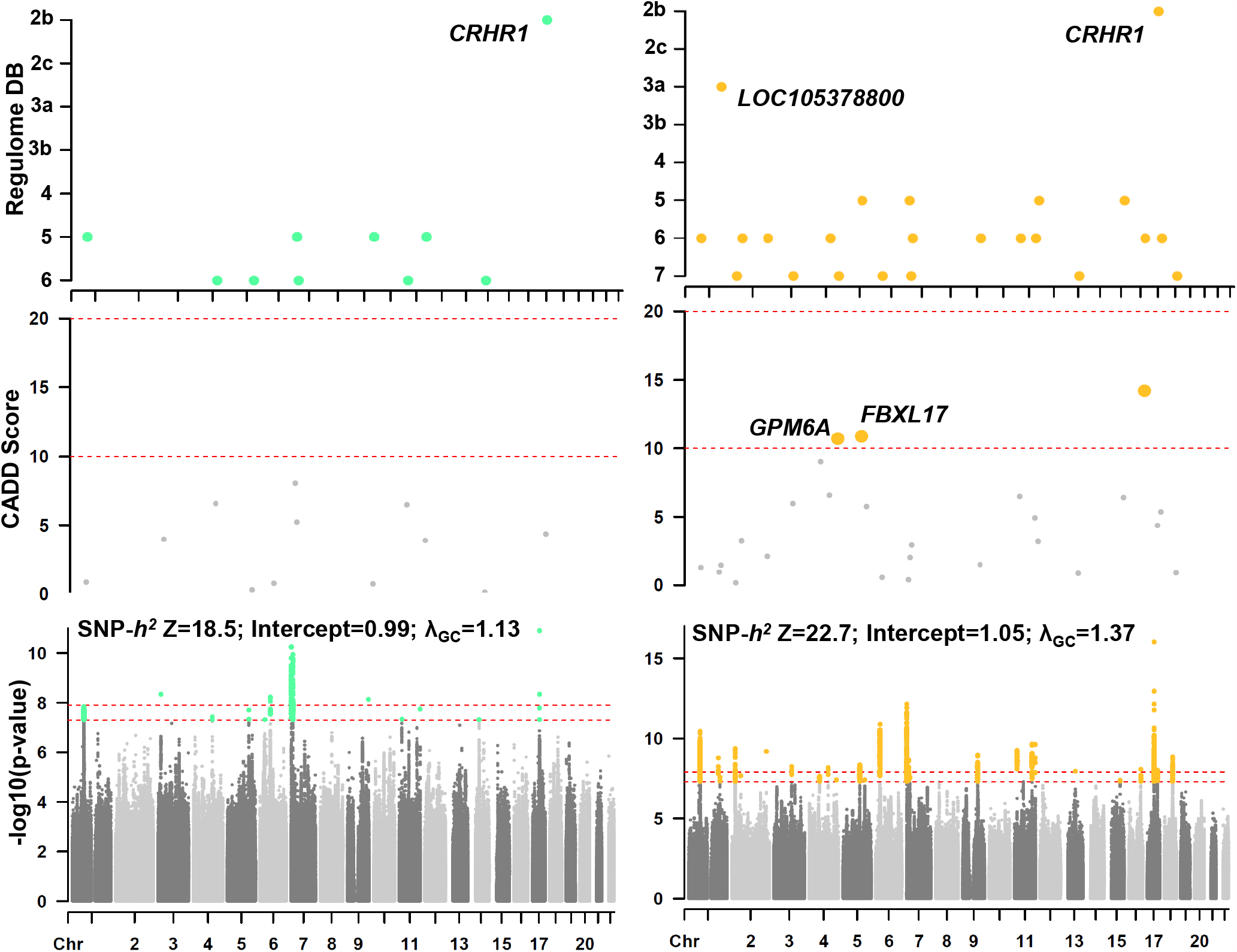
SNP annotation of GAD and PTSD GWAS. The bottom row shows Manhattan plots for each trait. Two horizontal dashed lines in each plot show the genome-wide significance threshold per phenotype (*P*<5×10^−8^) and study-wide (*P*_*adj*_<1.25×10^−8^). Above each Manhattan plot are Combined Annotation Dependent Depletion (CADD) scores and RegulomeDB scores for each genome-wide significant locus.

Out-sample PRS and functional annotation in the following sections were performed using the most powerful conditioned meta-analysis for each outcome (*i*.*e*., highest *h*^*2*^ z-score): meta-analysis of GAD using GAD-7 PheRS (*h*^*2*^=2.96%, s.e.=0.16, *P*=2.06×10^−76^) and meta-analysis of PTSD using PCL-6 PheRS (*h*^*2*^=4.08%, s.e.=0.18, *P*=8.86×10^−114^; Fig 3. and Supplementary Table 12).

### Out-Sample PRS

Out-sample PRS were calculated two ways. First, we evaluated overlap between the meta-analyses from this study with previous GWAS of anxiety and PTSD traits from FinnGen (KRA_PSY_ANXIETY N=15,770 cases and 161,129 controls; F5_PTSD N=781 cases and 161,390 controls), the PGC (PTSD v1 N=2,424 cases and 7,113 controls)^23^, and ANGST (N=17,310).^24^ At all *P*-value thresholds (*P*_*T*_), GAD and PTSD GWAS from this study predicted all out-sample GWAS (*P*<0.05; Supplementary Table 13). The maximum association for each trait was: GAD *versus* FinnGen KRA_PSY_ANXIETY (*R*^*2*^=0.029%, *P*_*T*_ =0.1, *P*_*T*_ =2.34×10^−13^) and PTSD *versus* PGC PTSD v1 (*R*^*2*^=0.006%, *P*_*T*_ =0.3, *P*=6.03×10^−4^).

Second, GAD and PTSD meta-analyses were used to predict GAD and PTSD symptoms in individual-level data from the Philadelphia Neurodevelopmental Cohort^25, 26^ and Yale-Penn^27, 28^ (Fig. 5). All PRS models were significant with at least one *P*_*T*_ (*P*<0.05) but GAD (*R*^*2*^=0.103%, *P*_*T*_=5×10^−8^, *P*=0.015) and PTSD meta-analyses (*R*^*2*^=0.874%, *P*_*T*_=1×10^−7^, *P*=1.57×10^−4^) best predicted corresponding symptoms in the PNC.

**Fig. 5.**
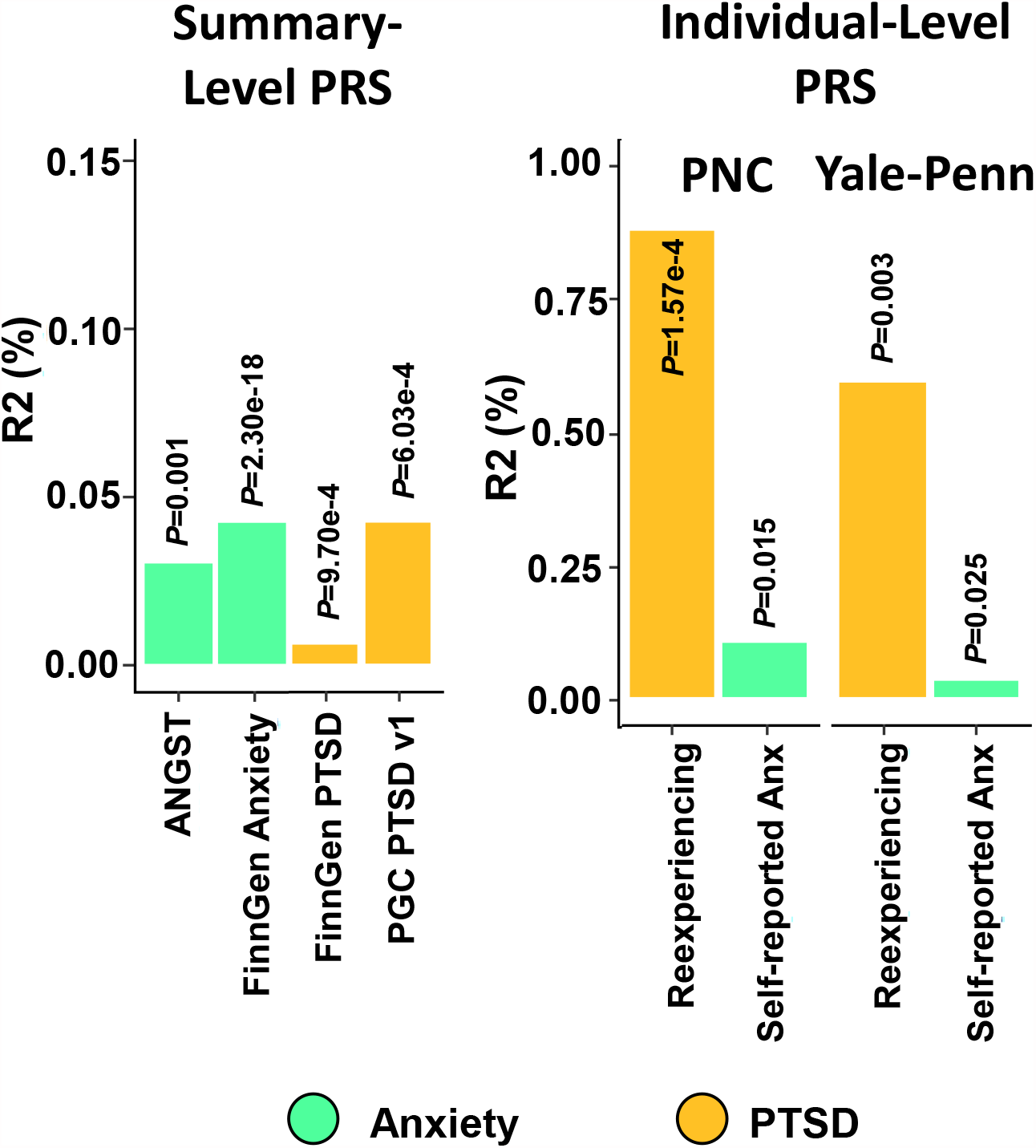
Out-sample polygenic prediction of relevant phenotypes. Maximum observed association (*R*^*2*^) between polygenic risk scores (PRS) for GAD and PTSD outcomes in this study and out-sample GAD and PTSD phenotypes from large consortia (ANGST, FinnGen, and PGC using summary-level PRS in PRSice v1.25) and individual-level cohorts informative for mental health outcomes (Philadelphia Neurodevelopmental Cohort (PNC) and Yale-Penn using PRSice v2).

### Prenatal Transcriptomic Enrichment

GWAS of GAD and PTSD were enriched for loci associated with multiple brain tissues (Supplementary Table 14). Brodmann Area 9 (BA9, a component of the dorsolateral prefrontal cortex) was enriched in both traits (GAD *β*=0.022, s.e.=0.007, *P*=9.07×10^−4^; PTSD *β*=0.030, s.e.=0.007, *P*=2.85×10^−5^). Each GAD and PTSD GWAS also was associated with transcriptomic profiles from the prenatal developmental stage (Fig. 6): GAD and late-mid prenatal tissue (*β*=0.041, s.e.=0.014, *P*=0.003); PTSD and late-mid prenatal tissue (*β*=0.042, s.e.=0.015, *P*=0.003; Supplementary Table 15). We attribute these enrichments to greater sample size and improved power relative to prior studies rather than multi-trait conditioning with *neuroticism score* as evidenced by detection of concordant enrichments in the unconditioned results (Supplementary Tables 14 and 15).

**Fig. 6.**
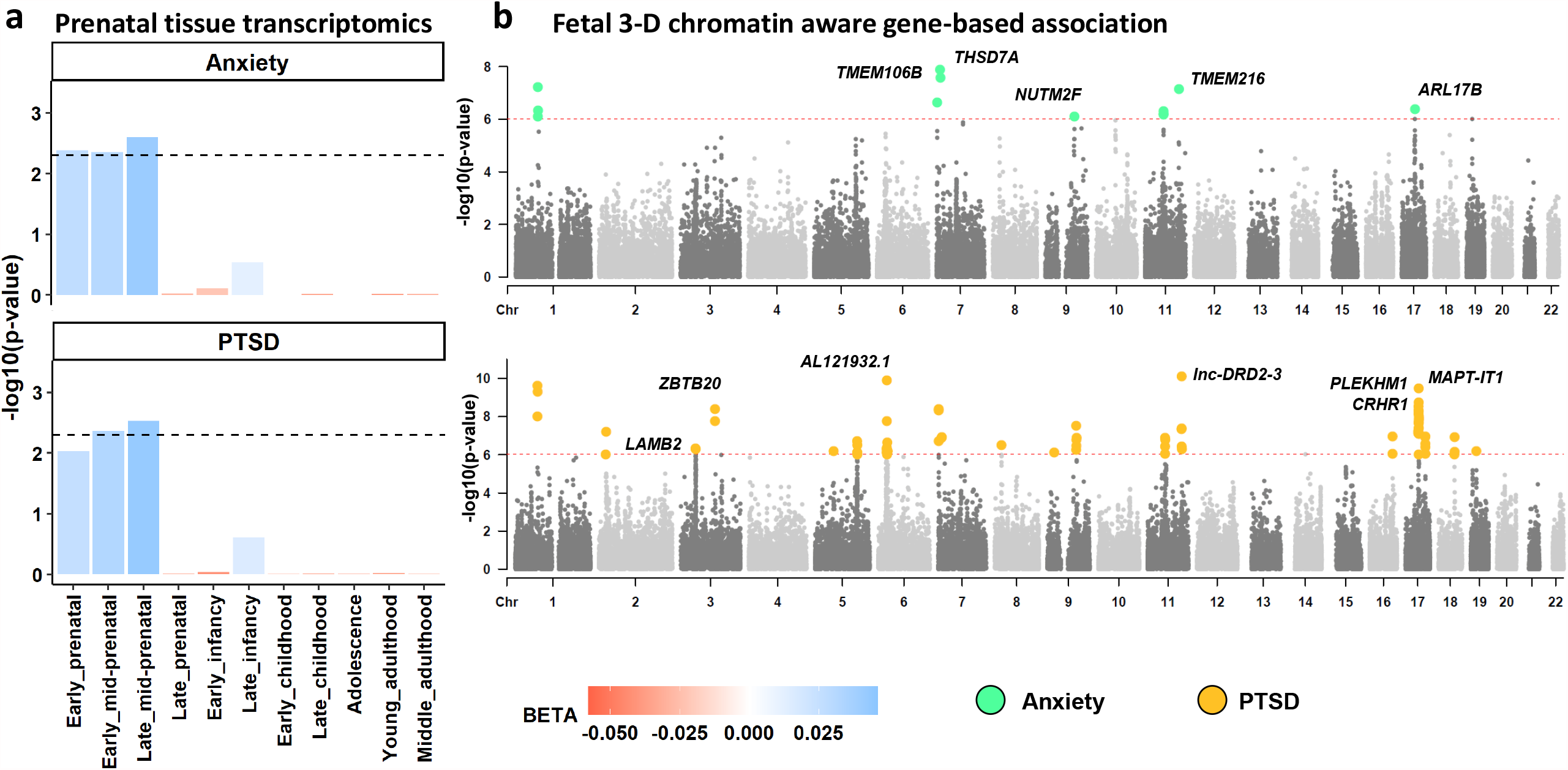
Prenatal transcriptomic signatures of GAD and PTSD outcomes. **a**, Enrichment of transcriptomic profiles from prenatal tissue based on BrainSpan 11 developmental stages. Each bar represents the results from one-sided tests for enrichment of a given transcriptomic profile. Effect size estimates (*β*) are color coded. Dashed horizontal lines indicate the significance threshold after multiple testing correction (FDR<0.05) across all tests. **b**, Manhattan plots of Hi-C coupled gene-based association studies of GAD and PTSD in fetal paracentral tissue. Each data point represents a single gene positionally aligned across each autosome. The height of each point along the y-axis indicates the significance of association between gene and phenotype with each colored data point indicating a significantly associated gene after analysis-wide multiple testing correction (*P*<9.43×10^−7^). A subset of genes are labeled and all genes are provided in Supplementary Table 16.

To investigate further the enrichment of prenatal tissue effects, we performed 3-D chromatin-aware gene-based association of GAD and PTSD GWAS using fetal brain tissue. We uncovered 135 GAD and 927 PTSD associated genes (*FDR*<0.05) in the context of fetal chromatin interactions including *CRHR1* (GAD *P*=1.54×10^−5^; PTSD *P*=1.75×10^−8^), *THSD7A* (GAD *P*=1.27×10^−8^; PTSD *P*=1.29×10^−7^), and *LAMB2* (GAD *P*=5.16×10^−5^; PTSD *P*=4.44×10^−7^). After study-wide multiple testing correction (*FDR*<0.05), 86 and 584 genes were associated with GAD and PTSD, respectively (Supplementary Table 16).

Consistent with tissue and 3-D chromatin analyses, cell-type enrichments reinforce the contribution of prenatal development in the liability for GAD and PTSD outcomes (Supplementary Table 17 and 18). GAD and PTSD GWAS were enriched for independent signals from post-conception prefrontal cortex neurons.^29^ In PTSD, two brain cell types had cross-dataset significant effects: GABAergic neurons from gestational week 26 (GW26) prefrontal cortex tissue (*β*=0.041, s.e.=0.012, *P*=2.64×10^−4^) and from the midbrains of 6-to-11-week-old embryos (*β*=0.246, s.e.=0.050, *P*=5.58×10^−7^). In GAD, the same two cell types were observed but did not survive multi-trait conditioning with *neuroticism*. Cross-data set analyses support partial independence of these signals with primary effects from 6-to-11-week-old midbrain neurons (proportional significance of midbrain GABAergic neurons given prefrontal cortex GABAergic neurons: PS_Mid_NbGaba,GW26_Gaba_ =0.700; PS_GW26_NbGaba,Mid_Gaba_=0.269).^19^

### Drug Targets and Repositioning

We investigated two means of drug targeting for the GAD and PTSD outcomes studied. First, we tested for *r*_*g*_ and evidence of latent causal effects (LCV) between GAD, PTSD, and medication use GWAS.^30, 31^ GAD and PTSD GWAS were most strongly genetically correlated with opioid use (GAD *r*_*g*_=0.530, s.e.=0.035, *P*=1.08×10^−50^; PTSD *r*_*g*_=0.603, s.e.=0.028, *P*=1.27×10^−100^) and antidepressant use (GAD *r*_*g*_=0.597, s.e.=0.041, *P*=1.94×10^−48^; PTSD *r*_*g*_=0.632, s.e.=0.035, *P*=3.23×10^−74^). There were no significant differences in medication use *r*_*g*_ due to multi-trait conditioning with *neuroticism* (Supplementary Table 19). We detected a single putative causal relationship (*FDR*<0.05) between use of vasodilators and GAD (genetic causality proportion=0.093, s.e.=0.285, *P*=1.04×10^−4^; Supplementary Table 20). Two-sample Mendelian randomization based on non-overlapping datasets (MVP GAD two-item questionnaire and UKB “use of vasodilators”) was insufficiently powered to confirm this causal hypothesis (Supplementary Results and Supplementary Table 21).

Second, we applied gene-ontology based drug repurposing. Gene-ontology enrichments were performed by mapping each LD-independent locus to a single gene to avoid redundant signals from nearby genes. Relative to the lead SNP in each LD-independent genomic risk locus we retained the nearest gene with the highest probability of loss of function intolerance (9 GAD and 17 PTSD genes; Supplementary Table 22) and detected 87 GAD and 28 PTSD gene-sets (FDR<0.05, *P*_*adj*_=4.55×10^−6^ based on 72,394 human gene sets,^32^ Supplementary Table 23). Forty-five drugs were nominally associated with both GWAS (Supplementary Table 24) highlighting compounds with evidence as treatment for anxiety disorders (*e*.*g*., dexpropranolol^33, 34^ GAD *P*=0.006, EScore=0.308; PTSD *P*=0.032, EScore=0.434). After multiple testing correction (FDR<0.05 applied per trait) we uncovered (i) upregulation of GAD gene-sets in the context of aminohippuric acid, a putative biomarker of depression and anxiety disorders (*P*=1.08×10^−5^, EScore=0.917).^35^

## Discussion

Extremely large cohorts are required to discover polygenic signals associated with anxiety^12, 24, 36, 37^ and PTSD.^4, 5, 23, 38^ Biobanks, such as UKB, offer a unique opportunity to boost sample size, power, and trait ascertainment homogeneity. In practice, studying mental health in the UKB is limited by the number of MHQ responders (only 31% of participants^8, 11, 12^) and the non-random missingness in the questionnaire participation. Due to the large proportion of missingness in the UKB MHQ, we aimed to maximize the sample size informative for GAD and PTSD by predicting quantitative and case-control phenotypes using elastic net regression in the UKB MHQ non-responders, then using the predicted traits in GWAS. We extend this prediction by using elastic net regression weights to calculate co-phenome PheRS.

We found that predicted GAD and PTSD outcomes and co-phenome PheRS reliably capture the genetic architecture of these traits of interest. Unsurprisingly, *neuroticism score* was a major correlate of each outcome and contributed substantial predictive power to the elastic net regression. We showed that in the context of income and internalizing spectrum psychopathologies, higher neuroticism scores were paradoxically associated with higher probability of participating in the MHQ. However, this effect appears to be driven by UKB participants with low-to-medium GAD-7 scores. While models of MHQ participation included GAD-7, PCL-6, and PHQ-9, all of which rely on the neuroticism score in their elastic net prediction, we captured independent effects such that accounting for GAD-7 changes the effect direction of *neuroticism* in a way that PCL-6 and PHQ-9 do not. This indicates that the change in the effect direction is not an artifact of co-phenome prediction and is also consistent with prior evidence of a two-factor model of *neuroticism*.^39^ The effect of neuroticism on MHQ participation may be due to liability of a subtype rather than neuroticism more broadly. Our data suggest that the interplay between *neuroticism* and GAD directly affect the MHQ participation bias. This could be due to the elevated anxiousness/tenseness elements rather than worry/vulnerability elements of *neuroticism*.^39^ *Neuroticism* is a transdiagnostic psychosocial factor, and this observation may indicate that (i) MHQ responders are characterized by less severe internalizing symptoms, (ii) MHQ responders may over-report the severity of these symptoms, and (iii) MHQ responders represent a subset of the UKB of greater socioeconomic status than non-responders.^39, 40^ We initially hypothesized that prior internalizing studies in the UKB were under powered due to sample size but the data reported here support a more refined hypothesis. Following previous evidence of a two-factor model of *neuroticism*, the depletion of a subtype (e.g., worry/vulnerability factor) in MHQ responders relative to non-responders may at least partially explain why previous GWAS of anxiety and PTSD traits in the UKB had only limited success and perhaps reinforces the relationship between *neuroticism* subtype, socioeconomic status, and mental health.^17, 39^ We leverage the higher *neuroticism scores* of MHQ non-responders to more than double the sample size upper limit for GWAS of GAD and PTSD outcomes while enriching the sample for individuals with objectively more severe symptoms. This procedure resulted in detection of more than twice the genomic risk loci associated with anxiety and PTSD relative than any of the previous studies.

Meta-analyses using PheRS were more powerful than meta-analyses using predicted quantitative traits. Though *h*^*2*^ differences between PheRS and quantitative outcomes were relatively small, we hypothesize that PheRSs capture slightly more accurate information about each trait because they are derived from tangential responses to questions not ascertained in the context of mental health (i.e., as part of the MHQ). In other words, studying the genetic liability of PheRS in combination with a directly ascertained quantitative symptom count may help reduce analytic noise associated with self-reported symptoms.^41^

Several approaches to risk locus functional annotation converged on fetal/prenatal biology. These findings are interesting given the childhood to mid-adult onset of internalizing disorders.^42^ We attribute this observation to the statistical power increases due to a larger sample size rather than multi-trait conditioning with *neuroticism*.^16, 17^ Consistent with previous studies,^43, 44^ the dorsolateral prefrontal cortex (DLPFC) was identified here as a tissue of interest for GAD and PTSD by GTEx tissue-wide analysis. We extended these observations to cell-type and 3-D chromatin interaction data to detect gestational week GABAergic neurons and several key genes of interest with significant effects in fetal brain tissue (GAD: *TMEM106B*; PTSD: *CRHR1, LAMB2*). In a prior single-cell RNA-seq study of the DLPFC (N=1,057 neurons), late gestational periods detected in our study were most enriched for genes related to axon guidance, neuron differentiation, axonogenesis, and regulation of neuron projection development.^29, 45^

We utilized the improved power of our meta-analyses to identify potential drug targets and/or drugs that induce anxiety and/or PTSD symptoms as adverse effects. The strongest genetic correlate of each trait was antidepressant use. We also detected a relationship between use of vasodilators and GAD using the LCV approach that could not be confirmed using a two-sample MR approach but has been detected by prior epidemiology research.^46^ The partial causal effect size of vasodilator use on GAD was small, so MR might be under-powered to detect this result considering (i) the requirement for non-overlapping samples and (ii) biases in LCV estimates in the presence of strong pleiotropic effects among highly polygenic traits.^31^ The discordance between these methods may reflect a causal relationship between GAD and vasodilator use that transcends a genetically-regulated molecular relationship (e.g., regulatory or proteomic elements).

The results from our study recapitulate and expand prior findings on the biology of GAD and PTSD but there are several key limitations to be considered. First, while capturing very similar genetic liability to GAD and PTSD, the elastic net-predicted phenotypes used in meta-analyses were weakly correlated with corresponding known GAD-7 and PCL-6 scores among UKB responders. Thus, there is no utility of these values for epidemiological studies of GAD or PTSD. However, they are valuable in genetic studies that aim to uncover biological mechanisms underlying psychopathology. Second, our study is restricted to participants of European ancestry and may/may not generalize to individuals of other ancestries. This lack of proven generalizability may be attributed both to genetic differences and documented variability in how racial and ethnic groups access and experience healthcare systems.^47^ In future work, our group and others will aim to recognize and reduce these health disparities using carefully tailored co-phenomes for these populations.^13^ Third, machine learning (i.e., elastic net regression) identifies patterns in data, not necessarily trait relationships. Thus, our analysis identified mathematically informative and biologically meaningful relationships with which to predict GAD and PTSD symptoms. However, these features, their predictive patterns, and the regression weights reported here may not translate outside the UK Biobank. Future studies will need to investigate how well PheRS created in one cohort may generalize to other cohorts. Finally, solutions to non-random missingness can be influenced by the proportion of missingness in a given dataset. Our data support the non-random nature of UKB MHQ missingness with respect to certain features of mental health (e.g., higher *neuroticism scores*) but this attribute of data missingness may not generalize to other biobanks. Future work on the generalizability of these findings will require detailed investigation of the type of missingness observed, its proportion relative to non-missing data, and how best to fill those gaps including the utility of other machine learning and/or imputation pipelines.

The elastic net predicted phenotypes derived here permit studies of GAD and PTSD in the whole UKB cohort. Our results provide one roadmap by which the community may improve sample size, and statistical power, for enhanced risk locus discovery in the context of incompletely ascertained traits in the UKB and other biobanks with limited mental health assessment. We use these data to present biological underpinnings uncovered from analysis of the largest GWAS meta-analysis of these traits to date.

## Online Methods

### Participants

The UKB is a large population-based cohort of over 502,000 participants between the ages of 37 and 73 at the time of recruitment. UKB assesses a wide range of factors in a generally healthy cohort including physical health, anthropometric measurements, circulating biomarkers, and sociodemographic characteristics. A subset of individuals (*N*=157,366) completed an ancillary online mental health questionnaire (MHQ)^11^ covering topics of self-reported mental health and well-being.

### GAD-7 Definition

GAD-7^48^ represents the summed score of seven questions from the UKB MHQ all of which are ranked by participants from 1=“Not at all,” 2=“Several days,” 3=“More than half the days,” to 4=“Nearly every day” with respect to how frequently they have been bothered by a given problem over the last two weeks. GAD-7 symptom items are Field ID 20505 “recent easy annoyance of irritability,” Field ID 20506 “recent feelings of nervousness or anxiety,” Field ID 20509 “recent inability to stop or control worrying,” Field ID 20512 “recent feelings of foreboding,” Field ID 20515 “recent trouble relaxing,” Field ID 20516 “recent restlessness,” and Field ID 20520 “recent worrying too much about different things.^11^” The mean GAD-7 score among UKB MHQ responders of European ancestry was 8.97 ± 3.09 (*N*=124,534). GAD-7 scores were stratified into current anxiety symptoms cases and controls using a cutoff threshold of GAD-7 ≧ 10 (*N*_*case*_ = 81,072; *N*_*control*_= 108,811).^12^

### PCL-6 Definition

PCL-6^49^ represents the summed score of six questions from the UKB MHQ. Five questions are ranked by participants from 0=“Not at all,” 1=“A little bit,” 2=“Moderately,” 3”Quite a bit,” to 4=“Extremely” with respect to a list of problems and complaints experienced in response to extremely stressful experiences over the past month. One questions (Field ID 20508) was ranked by participants from 1=“Not at all,” 2=“Several days,” 3=“More than half the days,” to 4=“Nearly every day” with respect to how frequently they have been bothered by a given problem over the last two weeks. PCL-6 symptom items are Field ID 20494 “felt irritable or had angry outbursts in past month,” Field ID 20495 “avoided activities or situations because of previous stressful experience in past month,” Field ID 20497 “repeated disturbing thoughts of stressful experience in past month,” Field ID 20498 “felt very upset when reminded of stressful experience in past month,” and Field ID 20508 “recent trouble concentrating on things.^11^” The mean PCL-6 score among UKB MHQ responders of European ancestry was 6.59±3.68 (*N*=126,219). PCL-6 scores were stratified into cases and controls using a cutoff threshold of PCL-6>13 (*N*_*case*_=3,663; *N*_*control*_=181,232).^4^

### PHQ-9 Definition

PHQ-9^48^ represents the summed score of nine items from the UKB MHQ the UKB MHQ all of which are ranked by participants from 1=“Not at all,” 2=“Several days,” 3=“More than half the days,” to 4=“Nearly every day” with respect to how frequently they have been bothered by a given problem over the last two weeks. PHQ-9 MHQ items are Field ID 20507 “recent feelings of inadequacy,” Field ID 20508 “recent trouble concentrating,” Field ID 20510 “recent feelings of depression,” Field ID 20511 “recent poor appetite or overeating,” Field ID 20513 “recent thoughts of suicide or self-harm,” Field OD 20514 “recent lack of interest or pleasure in doing things,” Field ID 20517 “trouble falling or staying asleep, or sleeping too much,” Field ID 20518 “recent changes in speed/amount of moving or speaking,” Field ID 20519 “recent feelings of tiredness or low energy.^11^” The mean PHQ-9 score among UKB Europeans who completed the MHQ was 11.73±3.67.

### Million Veteran Program Phenotypes

GAD-2 represents the summed score of self-report responses to two-questions regarding how bothered they felt by the following problems: (i) feeling nervous, anxious, or on edge and (ii) not being able to stop or control this worrying. Each question was ranked from 0=”Not at all” to 3=”Nearly every day” and was contextualized with feelings experienced over the two weeks prior to responding. The GAD-2 GWAS consisted of 175,163 participants of European ancestry producing an estimated GAD-2 *h*^*2*^=5.75%±0.42.^36^

PCL-Total (or PCL-17) is the summed score of 17 questions about the extent to which a participant has been affected by a given experience. Each question was ranked from 1=”Not at all” to 5=”Extremely” with respect to experiences over the past month. The PCL-Total GWAS consisted of 186,689 participants of European ancestry producing an estimated GAD-2 *h*^*2*^=8.84%±0.48.^5^

The GWAS of broad depression^3^ included 1,154,267 individuals from the MVP, 23andMe Inc., UKB, and FinnGen.^1, 50^ The MVP cohort included cases with at least one inpatient code or two outpatient codes for major depressive disorder and controls with no record or in- or outpatient codes for depression. Subjects with only one outpatient depression code were excluded. Eighteen ICD codes were considered for case inclusion. The *h*^*2*^ of broad depression in this large meta-analysis was 11.3%±0.38.

### First-Pass Feature Selection

The UKB contains thousands of potentially informative phenotypes for predicting a given outcome. We narrowed our focus to a subset of these with which to perform elastic net regression (see Elastic Net Regression Parameter Optimization). We selected phenotypes with more than 200,000 responses, not part of the MHQ, and lacking highly dimensional structure (e.g., ICD-9/10 codes, medication endorsements (Field ID 20003)), and those attributes available through special requests (e.g., greenspace (Field ID 24500) and water percentages (Field ID 24502)). The final feature set included 772 phenotypes.

### Elastic Net Regression Parameter Optimization

To focus the selection of co-phenome features, we tested for phenome-wide Spearman correlations between GAD-7 and PCL-6 and all 772 features in the UKB MHQ responders of European ancestry (total *N*=132,016). Removing related individuals was done per trait such that the member of each related pair with the larger symptom count could be retained. Using the magnitude of rho for all nominally significant features of the phenome, we performed elastic net regression three ways to determine optimal model parameters with which to predict GAD-7 and PCL-6 in the UKB sample lacking MHQ data. Three training and test proportions (25%_train_| 75%_test_, 50%_train_| 50%_test_, and 75%_train_| 25%_test_) were tested for four thresholds of rho used to include features in the regression: *ρ*>0.3, >0.25, >0.2, >0.15. Elastic net regression was performed with the `glmnet` R package^51^ for each parameter setting using standardized 50-fold cross validation using the best-fit penalizing parameter lambda. Parameter combination success was determined by comparing predicted GAD-7 and PCL-6 to MHQ-derived GAD-7 and PCL-6 using Spearman correlation. Test results are described in Supplementary Table 2. Using the optimal feature inclusion settings, feature weights (elastic net *β*) were extracted with which to calculate co-phenome risk scores (see **Creation and Reliability of Co-Phenome Risk Score**).

### Creation and Reliability of Co-Phenome Risk Scores

Phenotype risk scores were calculated as described previously.^13^ Briefly, we calculated GAD and PTSD risk scores as the weighted sum of the co-phenome. Each phenotype was weighted by the effect size obtained from 50-fold cross validation elastic net regression. GAD and PTSD co-phenome 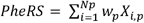, where *N*_*p*_ is the number of comorbid phenotypes determined by Spearman 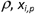 is set to 0 if the trait response was coded as missing, “prefer not to answer,” or a comparable derivative indicating a non-answer to the UKB question and *w*_*p*_ is the effect size (*β*) obtained from elastic net regression.

We determined the reliability of predicted GAD and PTSD outcomes and co-phenome PheRS several ways. First, within the MHQ responders and MHQ non-responders, we correlated quantitative outcomes, co-phenome PheRS, and categorical diagnoses. Second, based on GWAS of each GAD and PTSD outcomes, we performed genetic correlation within and between MHQ responders and MHQ non-responders. As a positive control with positive *r*_*g*_, we included the GWAS of neuroticism from the Social Science Genetic Association Consortium (SSGAC);^16^ as a positive control with negative *r*_*g*_, we included the GWAS of subjective well-being from SSGAC.^16^ Third, we calculated polygenic risk scores for each unrelated European-ancestry participant in the UKB using GAD-2 (GWAS *N*=199,611)^36^ and PTSD PCL-17 (GWAS *N*=186,689)^5^ from the Million Veteran Program (MVP). To our knowledge, these GWAS represent the largest and most powerful genetic assessments of GAD and PTSD outcomes with no known overlap with UKB. PRS were calculated with PRSice v2^52^ with the following clumping parameters to select linkage disequilibrium independent variants: *r*^*2*^=0.001, *P*=1, in 10,000-kb windows. The range of GAD SNPs included in the PRS at minimum and maximum *P*-values thresholds (*P*_*T*_) was 5 (*P*_*T*_ =5×10^−8^) to 4,060 (*P*_*T*_ =1); the range of PTSD SNPs included in the PRS at minimum and maximum *P*-value thresholds: 14 (*P*_*T*_ =5×10^- 8^) to 10,264 (*P*_*T*_ =1). Relationships between PRS, co-phenome PheRS, quantitative outcomes, and case-control diagnoses were tested using age, sex, age×sex, and the first ten within-ancestry principal components as covariates.

### Modeling MHQ Participation

We build a logistic regression model of MHQ participation using UKB participants who completed the online MHQ (Supplementary Table 5a). To evaluate the effects of each predictor, we estimated predicted probabilities of MHQ participation using the R package `effects`.^53^ In R, the top two predictors of MHQ participation were sampled 100 times at each stratum of neuroticism score (range 0-12). Multicollinearity among model predictors was tested using `mctest`.^54^ Briefly, `mctest` computes overall (i.e., model-level) and individual (i.e., variable-level) multicollinearity diagnostics. The package therefore permits the identification of which predictors are the source of collinearity.

### GWAS of GAD and PTSD Co-Phenome Risk Scores and Predicted Outcomes

Genotyping and imputation of the UKB cohort has been previously described.^8^ Briefly, UKB participants were genotyped using custom Axiom array with marker content chosen to capture genome-wide genetic variation and short insertion/deletions, including coding variants across a range of minor allele frequencies and markers providing good coverage for imputation in European ancestry populations. UKB was imputed using the Haplotype Reference Consortium reference panel.

Genome-wide association studies (GWAS) of GAD and PTSD were performed in two ways: (i) in the subset of UKB MHQ responders and (ii) in the UKB MHQ non-responders using predicted GAD and PTSD outcomes. For this study, all GWAS were restricted to UKB participants of European ancestry as identified by a two-stage ancestry assignment and pruning procedure from the Pan-UKB. Briefly, PCA was performed on unrelated individuals from a combined reference panel from the 1000 Genomes Project and Human Genome Diversity Project. A random forest classifier was trained to assign continental ancestry assignments based on six principal components. Unknowns from the UKB were assigned to an ancestry group if their random forest probability was >50%. This list was further refined to remove ancestry outliers based on within-population PC distances from 3-5 centroids. Detailed description of these methods is provided at https://pan.ukbb.broadinstitute.org/docs/technical-overview.

Linear regression was performed in PLINK 2.0 using SNPs with imputation INFO scores > 0.8, minor allele frequencies > 0.01, missingness < 0.05, and Hardy-Weinberg equilibrium *P*-values > 1×10^−10^. We included age, sex, age×sex, and the first ten within-ancestry principal components as covariates in each GWAS. GWAS of each trait were meta-analyzed with the MVP counterpart GWAS using METAL.^55^ Per meta-analyzed GWAS, we applied a genome-wide significance threshold of 5×10^−8^. To account for multiple testing correction, we also considered a study-wide significance threshold of 1.25×10^−8^ = 0.05/1,000,000 LD independent SNPs in EUR/2 meta-analyses/2 internalizing traits.

### Locus Fine-mapping

We fine-mapped the association statistics of four phenotypes (GAD and PTSD phenotype risk scores and two quantitative outcomes) for a 1MB region around the LD-independent significant SNPs as identified by FUMA (FUnctional Mapping and Annotation (FUMA v1.6a; *r*^*2*^=0.6 in 2-kb windows; 1-kb window on each side^19^). Each region for the respective association was fine-mapped to determine the 95% credible set using `susieR`^56^ with at most 10 causal variants. The credible set reports whether each identified variant is among a set of most likely causal variants and the marginal posterior inclusion probability (PIP) for causal set membership. PIPs range from 0 to 1 with values closer to 1 indicating greater causal probability.

### LDSC and SNP-based Heritability

Observed-scale heritability was calculated for each GWAS using Linkage Disequilibrium Score Regression (LDSC)^30^ with the 1000 Genomes Project European ancestry reference LD panel. LDSC also was used to calculate (i) genetic correlation between GWAS in MHQ responders and MHQ non-responders, (ii) genetic correlation between GAD and PTSD outcome GWAS from the UKB and MVP, and (iii) genetic correlation between medication use traits from UKB and meta-analyzed GAD and PTSD outcomes (i.e., UKB + MVP). Liability-scale *h*^*2*^ estimates were generated using the sample case prevalence (Supplementary Table 6) and population-prevalence as following: GAD = 16%,^57^ PTSD = 7%,^58^ MDD = 20%.^59^

### Functional Annotation

GAD and PTSD outcome liability loci were positionally mapped with Multi-marker Analysis of GenoMic Annotation (MAGMA v1.08) implemented in FUMA v1.6a^60^ using 2-kb positional mapping around each lead SNP.^19^ Linkage disequilibrium independent genomic risk loci are defined by their lead SNP (*P*<5×10-8) and all surrounding SNPs with *r*^*2*^>0.6 with the lead SNP.

Enrichment of tissue transcriptomic profiles was tested relative to Genotype-Tissue Expression (GTEx v8^61^) 53 tissue types and the BrainSpan Atlas of the Developing Human Brain^62^ 29 brain ages ranging from 8 weeks post-conception to 40 years old and 11 general developmental stages of the brain ranging from early prenatal to middle adulthood.

Cell-type transcriptomic profile enrichments were performed using 13 human-specific transcriptomic profile datasets related to the brain: PsychENCODE_Developmental, PsychENCODE_Adult, Allen_Human_LGN_level1, Allen_Human_LGN_level2, Allen_Human_MTG_level1, Allen_Human_MTG_level2, DroNc_Human_Hippocampus, GSE104276_Human_Prefrontal_cortex_all_ages, GSE104276_Human_prefrontal_cortex_per_ages, GSE67835_Human_Cortex, GSE67835_Human_Cortex_woFetal, Linnarson_GSE101601_Human_Temporal_cortex and Linnarson_GSE76381_Human_Midbrain. Cell-type transcriptomic profiles were assessed in three ways as per the FUMA analysis pipeline: (1) enrichment of cell-type transcriptomic profiles within each selected dataset, (2) within-dataset conditionally independent cell-type transcriptomic profile enrichments and (3) across-dataset cell-type transcriptomic profile enrichments.^19^

For analyses within datasets, cell type conditional significance is evaluated per dataset against *P*-values for all other cell types in that dataset. The output from these analyses identify cell types within a dataset whose transcriptomic profiles are enriched in a given GWAS independently of the signal from all other cell-type transcriptomic profiles in the same dataset.

Cell types from the same dataset may be similar. Using within-dataset significant cell types identified above, conditional analysis identifies cell-type transcriptomic profiles enriched in a given GWAS independent of all other cell types from the selected dataset by setting thresholds for proportional significance (PS) of the conditional *P*-value of a cell type relative to the marginal *P*-value. PS and conditional independence of cell-type pairs indicate that enrichment of these cell types in a given

GWAS is driven by independent genetic signals. The PS of cell type *a* given cell type *b* is 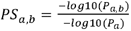 where *P*_*a*_ is the marginal *P*-value for cell-type a using the baseline model of cell-type enrichment (*Z*_*a*_ = *β*_0_ + *E*_*a*_*β*_*Ea*_ + *Aβ*_*A*_ + *Bβ*_*B*_ + *ϵ*) and Pa,b is the conditional *P*-value for cell-type *a* given the effects of cell-type *b* (*Z*_*a,b*_ = *β*_0_ + *E*_*a*_*β*_*Ea*_ + *E*_*b*_*β*_*B*_ + *Aβ*_*A*_ + *Bβ*_*B*_ + *E*). For a given pair of cell types, PS_*a,b*_ ≥ 0.8 and PS_*b,a*_ ≥ 0.8 indicates independent genetic signals for cell types *a* and *b*. Interpretation of additional PS thresholds for each cell type in a given pair can be seen in detail online (https://fuma.ctglab.nl/tutorial#celltype) or in Watanabe et al.^19^

Conditional analyses may also be applied across datasets using all possible combination of within-data set independent signals. Similar to conditioning within a dataset, the cross-data conditional P-value of a cell type is compared to the cross dataset marginal *P*-value for the same cell type, resulting in a cross-dataset PS value.

### Medication Use GWAS

Twenty-three GWAS of medication use were evaluated for genetic overlap and causal relationships with GAD and PTSD. Each medication use GWAS^63^ tested for association between approximately 7 million SNPs and the endorsement of medication in a category (e.g., diuretics, opioids, antidepressants, etc.) in more than 320,000 European ancestry participants from the UKB. Data may be accessed here: https://cnsgenomics.com/content/data.

The SNP-*h*^*2*^ of all 23 medication use GWAS were significantly different from zero; however, one (L04-Immunosuppressants) was insufficiently powered for genetic correlation (*h*^*2*^ z-score=3.28) as recommended by the LDSC developers. Twenty-two suitably powered medication use GWAS (*h*^*2*^ z-score>4), were tested for genetic correlation with each trait based on the 1000 Genomes Project LD reference using LDSC.

Latent causal variable (LCV^31^) analysis infers genetic causal relationships between trait pairs using SNP effect size estimates or Z-scores from GWAS summary association data. The LCV model assumptions are notably weaker than traditional Mendelian randomization assumptions. First, LCV assumes that the distribution of effect sizes for a given trait pair represents one distribution of proportional effects in both traits and a second distribution of effects specific to the outcome trait. In other words, LCV assumes that symmetry in shared genetic architectures between traits arises from the action of a latent genetic component rather than a non-genetic confounder commonly elucidated by MR. Second, LCV assumes a single latent factor mediating trait relationships; however, when more than one latent factor was simulated, LCV was unlikely to detect a causal effect. There are no assumptions of parametric effect size distributions under the LCV model, but LCV is less well powered for highly polygenic traits.

LCV modelling was implemented in R using the 1000 Genomes Project phase 3 European reference panel. GWAS summary data were filtered to include only SNPs with minor allele frequencies greater than 5% and the major histocompatibility region was removed due to its complex LD structure. LCV model output distinguishes whether genetic correlations support genetic causation and the degree to which (that is, the gĉp) genetic risk for trait 1 is causal for trait 2. LCV gĉp estimates range from 0 to 1 with values closer to 1 indicating full causality between two traits. Gĉp estimates were only interpreted for trait pairs where both traits exhibit LCV-calculated *h*^*2*^ Z-scores ≥7.

### Drug Repurposing

Drug repurposing was performed using the Gene2drug computational tool.^64^ Gene2drug applies pathway-set enrichment analysis to a group of gene-sets to reveal pathways of genes up- or down-regulated by a drug based on gene expression profiles from ConnectivityMap.^65^ Given a set of pathways or gene-sets, Gene2drug reports a *P*-value for the Kolmogorov-Smirnov statistic. Each drug is assigned an enrichment score (“EScore”) to describe the magnitude and direction of dysregulation with positive EScores indicating upregulation and negative EScores indicating downregulation.

To select Gene Ontology terms as standard input for Drug2gene, we positionally mapped the lead SNP from each genomic risk locus to its nearest gene. When more than one gene mapped to a lead SNP, we prioritized retention of the gene with the greatest probability of loss of function intolerance. This list of genes was submitted to ShinyGO.^32^ We extracted all significant GO terms after multiple testing correction (FDR<0.05, *P*_*adj*_=4.55×10^−6^ based on 72,394 human gene sets) as input for Gene2drug.

### External Cohort Polygenic Risk Scoring

#### Yale-Penn

participants were collected from five study sites in the eastern United States for studies of the genetics of substance dependence and anxiety traits using the Semi-Structured Assessment for Drug Dependence and Alcoholism (SSADDA).^27, 28^ Participants provided written informed consent through a protocol approved by the institutional review board at each participating site (Yale Human Research Protection Program (protocols 9809010515, 0102012183, and 9010005841), University of Pennsylvania Institutional Review Board, University of Connecticut Health Center Institutional Review Board, Medical University of South Carolina Institutional Review Board for Human Research, and the McLean Hospital Institutional Review Board).

#### The Philadelphia Neurodevelopmental Cohort (PNC)^25, 26^

consists of youths aged 8-21. PNC was designed to study the genomics of complex pediatric disorders but is not enriched for any specific disorder; the cohort is considered generally healthy. All participants underwent clinical assessment, including a neuropsychiatric structured interview and review of electronic medical records. They were also administered a neuroscience based computerized neurocognitive battery (CNB) and a subsample underwent neuroimaging. Clinical testing for each participant included (1) GOASSES (a modified version of the Kiddie-Schedule for Affective Disorders and Schizophrenia), demographic and medical history, Global Assessment of Functioning, and general interviewer observations, (2) a psychopathology symptom and criterion-related assessment of mood disorders, anxiety disorders, behavioral disorders, psychosis spectrum, eating disorders, suicidal thinking and behavior, and treatment history, and (3) an abbreviated form of the Family Interview for Genetics Studies to assess major domains of psychopathology in the proband’s first-degree relatives. A neurocognitive battery was performed for each PNC proband to measure accuracy and speed of executive control functions, episodic memory, social cognition, and sensorimotor and motor speed. For a complete list of neurodevelopmental domains, see https://www.ncbi.nlm.nih.gov/projects/gap/cgi-bin/study.cgi?study_id=phs000607.v3.p2.

We calculated PRS for each unrelated European participant of the Yale-Penn and PNC, for each GAD and PTSD symptoms: (i) reexperiencing (*N*_*PNC*_=2,796 (18% cases); *N*_*Yale-Penn*_=1,117 (84% cases)) and self-reported anxiousness (*N*_*PNC*_=3,053 (58% cases); *N*_*Yale-Penn*_=3,416 (25% cases)). Per cohort, European ancestry was defined by clustering principal components with the 1000 Genomes Project European reference population using the --cluster flag in plink. PRS were calculated in PRSice v2^52^ with the following clumping parameters to select linkage disequilibrium independent variants overlapping between this study’s meta-analysis and the SNP arrays for each cohort: *r*^*2*^=0.001, *P*=1, in 10,000-kb windows. In Yale-Penn, we used between 1,546-1,570 SNPs LD-independent SNPs; in PNC, we used between 1,501-1,514 LD-independent SNPs. Association between PRS and internalizing symptom included age, sex, age×sex, and the first ten within-ancestry principal components as covariates. We tested 11 *P*-value thresholds for SNP inclusion: 5×10^−8^, 1×10^−7^, 1×10^−6^, 1×10^−5^, 1×10^−4^, 0.001, 0.05, 0.1, 0.3, 0.5, 1.

GWAS summary association data were retrieved from the Psychiatric Genomics Consortium,^23^ Anxiety NeuroGenetics STudy (ANGST^37^), and the FinnGen initiative (Release 4; http://r4.finngen.fi/). With PRSice v1.25^66^ we performed a strict LD-clumping using *r*^*2*^=0.001, *P*=1, in 10,000-kb windows and testing 11 *P*-value thresholds: 5×10^−8^, 1×10^−7^, 1×10^−6^, 1×10^−5^, 1×10^−4^, 0.001, 0.05, 0.1, 0.3, 0.5, 1.

### Mendelian Randomization

We performed Mendelian randomization to test the relationship between vasodilator use and GAD using non-overlapping GWAS summary association data from Levey, et al. (GAD-2 in the MVP^36^) and Wu, et al. (vasodilator use in the UKB^63^). We performed two analyses in a bidirectional manner (e.g., GAD-2➔ vasodilator use and vasodilator use➔GAD-2): (i) using LD-independent genome-wide significant variants from the exposure GWAS (*P*<5×10^−8^) and (ii) using all LD-independent variants from the exposure GWAS. Similar to prior studies,^67, 68, 69^ we attempted a third MR instrument selection procedure using variants associated with the exposure at a *P*-value threshold producing the greatest polygenic prediction of the outcome (i.e., derived from PRS between exposure and outcome); however, this approach identified genome-wide significant threshold (*P*<5×10^−8^) as the most suitable threshold to test vasodilator➔GAD-2.

MR relies on three assumptions about the genetic instrument: (i) SNPs are associated with the exposure variable, (ii) SNPs are not associated with confounding factors linking the exposure to the outcome, and (iii) SNPs are associated with the outcome only through its association with the exposure. Using the R package `TwoSampleMR`, we tested different MR methods: random-effect inverse variance weighted (IVW), MR-Egger, weighted median, simple mode, and weighted mode.^70, 71, 72, 73^ These different approaches have different sensitivities with respect to different causal scenarios. We also conducted sensitivity tests for the presence of horizontal pleiotropy in the genetic instrument (MR-Egger regression intercept and MR-PRESSO (Pleiotropy RESidual Sum and Outlier) global test) and to investigate the heterogeneity of variants.^74, 75^ FDR multiple testing correction was applied to account for the number of MR tests performed.

## Supporting information

Supplementary Results

## Data Availability

All data used to generate figures for this study are provided as Supplementary Material. Elastic net weights are provided as Supplementary Material. GWAS summary data are accessible at 10.5281/zenodo.4767570. This research has been conducted using the UK Biobank Resource (application reference no. 58146) and is available to bona fide researchers through approved access. Out-sample polygenic risk scoring utilized the Yale-Penn cohort (dbGaP Study Accession: phs000425.v1.p1) and the Philadelphia Neurodevelopmental Cohort (dbGaP Study Accession: phs000607.v3.p2). The dbGAP data used herein is available for approved access download from dbGAP data request portal.

## Code availability

Previously developed pipelines were used to produce the results for this study. No custom code was developed to generate the data used to draw any of our conclusions.

## Acknowledgements

This research has been conducted using the UK Biobank Resource (application reference no. 58146). The authors thank the research participants and employees of the UK Biobank for making this work possible. This study was supported by National Institutes of Health (R21 DC018098, R21 DA047527, R33 DA047527, and F32 MH122058) and a Faculty Scholar Award from the Seaver Foundation: “Analytical Genomics of Vulnerable Populations.” The funders had no role in study design, data collection and analysis, decision to publish, or preparation of the manuscript.

## Author Contributions

F.R.W and R.P. conceived the study design; F.R.W. performed phenotype prediction, demographic comparisons, multi-trait conditioning, genome-wide association study meta-analyses, causal inference analysis, functional annotation, and PRS in the PNC; G.A.P. performed fine mapping; J.D.D. assisted with multi-trait conditioning; F.D.A. performed drug repurposing analyses; D.K. performed genetic correlation; B.C.M. performed GWAS statistics-level PRS; F.R.W., G.A.P., J.D.D., D.S.L., D.F.L, M.B.S., H.R.K., K.C.K., J.G., L.M.H., and R.P. contributed to data interpretation; F.R.W., G.A.P. and R.P. contributed to data visualization and presentation; F.R.W. drafted the original manuscript. All authors critically evaluated and revised the manuscript.

