## Supplementary Results for "Comorbid-phenome prediction and phenotype risk scores enhance gene discovery for generalized anxiety disorder and posttraumatic stress disorder"

**Multicollinearity in MHQ Participation Biases**

To verify the robustness of the observation that MHQ participation is, in part, a product of independent effects of “*neuroticism score*” and GAD, but not PTSD, depression or other variables evaluated herein, we tested for multicollinearity among independent variables in the model (Supplementary Table 5).

As there is no single “best” statistic for the detection of multicollinearity, we used the mctest R package to test for overall model indices of multicollinearity using multiple measures:^53^ Determinant |X'X| (an estimate of multivariable matrix relationships), Farrar-Glauber (F-G) Chi-squared statistic (a formal test for departure from matrix orthogonality) , *Red* indicator (a measure of variable redundancy in a given data frame), inverse |X'X| matrix relationships (an indicator of least squares computing errors), and Theil’s method which considers the pairwise correlation between dependent and each independent measure. The only significant overall measure of multicollinearity was observed with the F-G statistic (Chi-Squared = 1,036,100). The remaining indicators did not detect evidence of multicollinearity in the model.

Next, we evaluated which features contribute to multicollinearity as estimated by the F-G test. With very large sample sizes, it is unsurprising to detect large F-statistics per variable (Supplementary Table S5). By variance inflation factors (VIF), GAD and neuroticism score are slightly more inflated than the remaining variables suggesting possible multicollinearity. The exact value of VIF that indicates substantial multicollinearity is not standardized. It is generally recommended that a VIF > 10 indicates high correlation and high multicollinearity while some analyses suggest VIF > 2.5 is cause for concern.^72^ Because the correlation between GAD and neuroticism score is modest (*ρ*=0.644), and the VIF for GAD (2.46) and neuroticism score (2.57) do not surpass even the most conservative multicollinearity criteria, we rule out the likelihood of any appreciable multicollinearity affecting our model of MHQ participation.

**Drug Targets and Repositioning**

To test the putative causal relationship between vasodilator use and GAD-2 we performed MR two ways: (i) using GWS variants in the exposure and (ii) using all LD-independent variants in the exposure. To avoid sample overlap, a known confounder of MR, we used GWAS summary association data from Levey*, et al.* regarding GAD-2^35^ and from Wu, et al. regarding vasodilator use.^59^ Unless otherwise noted, inverse variance weighted (IVW) estimates are reported and all estimates, including the robust adjusted profile estimate accounting for weak genetic instruments, are provided in Supplementary Table 21).

*Vasodilator use* 🡪 *GAD-2* Three SNPs from the vasodilator use GWAS meet genome-wide significance (*P*<5x10^-8^). One SNP (rs1333047) is palindromic with intermediate allele frequencies and was removed; a second SNP (rs140570886) is not found in the MVP GAD-2 GWAS and we failed to identify a suitable LD-proxy within 1-kb up or downstream of the target SNP (LD *R^2^* ranged from 0.0009 (rs6793969) to 0.0058 (rs9457946)). We did not detect a significant causal effect (*P*=0.423, IVW *β*=0.030, s.e.=0.038) and because only a single variant was used, horizontal pleiotropy could not be assessed. To investigate whether this non-significant effect was due to a lack of power, we tested vasodilator use genetic instrument based on genome-wide LD-independent variants (*N*=2,628 SNPs). Tests with all LD-independent variants were influenced by heterogeneity (*P*=8.65x10^-6^, *Q*=2949.1) and horizontal pleiotropy (*P*=0.029, Egger intercept=0.001, s.e.=5.24x10^-4^). These effects could not be removed from the causal relationship even after eliminating outliers (60% confidence interval; N=2,132 SNPs) based on MR-PRESSO observed residual sum of squares and per-SNP effects (heterogeneity: *P*=1, *Q*=1429.8; horizontal pleiotropy: *P*=0.017, Egger intercept=0.001, s.e.=5.26x10^-4^; *P*=0.058, IVW *β*=0.005, s.e.=0.003).

### GAD-2 🡪 vasodilator use Using a subset of GAD-2 SNPs with P<5x10^-8^ (N=5 SNPs) and no evidence of heterogeneity (P=1.58x10^-4^, Q=2.13), there was no significant effect of GAD-2 on vasodilator use (P=0.187, IVW β=0.573, s.e.=0.427). Too few SNPs were used to test for horizontal pleiotropy. To investigate whether this non-significant effect was due to a lack of power, we tested a vasodilator use genetic instrument based on genome-wide LD-independent variants (N=1,8657 SNPs). We detected a nominally significant positive effect of GAD-2 of vasodilator use (P=0.033, IVW β=0.059, s.e.=0.028) in the absence of heterogeneity (P=0.325, Q=1883.1) and horizontal pleiotropy (P=0.517, Egger intercept=-0.001, s.e.=0.002) among the genetic instruments. This result did not survive multiple testing correction.

### Supplementary Figures

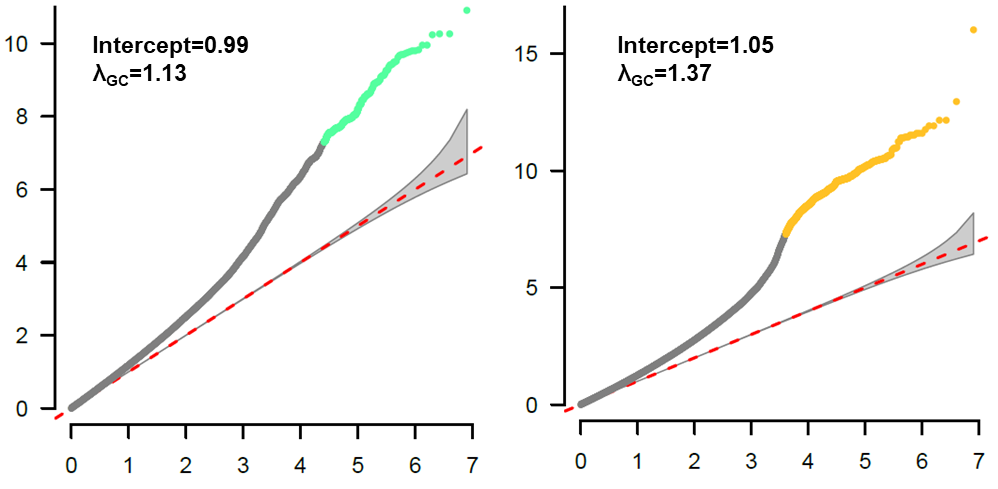

**Supplementary Fig. 1 |** Quantile-quantile plots of meta-analyzed GWAS of GAD (green) and PTSD (yellow).

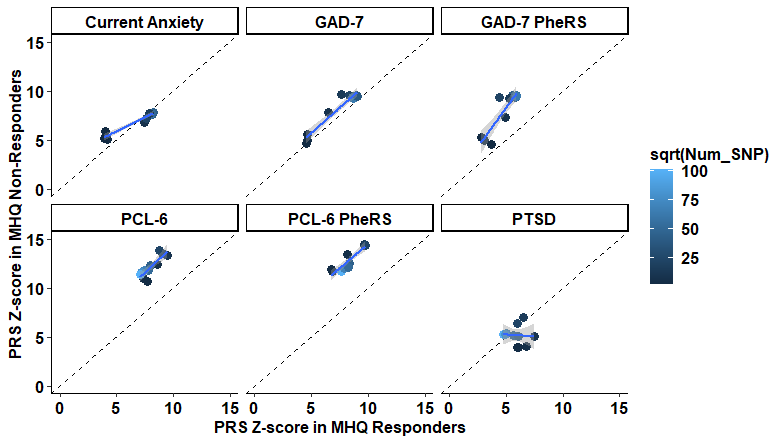

**Supplementary Fig. 2 |** Relationship between polygenic risk scores (PRS) from the Million Veteran Program generalized anxiety disorder (MVP GAD-2 predicting UKB current anxiety, GAD-7, and GAD-7-PheRS) and posttraumatic stress disorder (MVP PTSD Checklist 17-item score predicting UKB PTSD, PCL-6, and PCL-6-PheRS) in Mental Health Questionnaire responders and non-responders.

### Supplementary Tables

**Supplementary Table 1.** Significant (FDR<0.05 per internalizing phenotype) Spearman correlation between UK Biobank phenotypes and quantitative psychiatric outcomes in the UK Biobank participants who completed the Mental Health Questionnaire. GAD-7 = generalized anxiety disorder 7-item symptom count; PCL-6 = PTSD Checklist 6-item symptom count; PHQ-9 = depression 9-item symptom count.

**Supplementary Table 2.** Results of elastic net regression parameter tuning and selection. Parameters in red were selected to calculate feature weights for co-phenome risk scores.

**Supplementary Table 3.** Elastic net regression weights calculated for each internalizing co-morbid phenotype in the UK Biobank.

**Supplementary Table 4.** Spearman correlation and 95% confidence intervals for all internalizing trait pairs (A) and either *T*-statistic (quantitative outcomes include symptom counts and phenotype risk scores (PheRS)) or goodness of fit test statistics (case-control outcomes) for each outcome between UK Biobank participants who did and did not complete the Mental Health Questionnaire (MHQ).

**Supplementary Table 5.** Demographic predictors of Mental Health Questionnaire (MHQ) participation. In (A) independent predictors of MHQ participation were detected. In b (full model), c (full model minus GAD-7), d (full model minus PCL-6), e (full model minus PHQ-9) we show the flipped effects of neuroticism scores on MHQ participation (highlighted in yellow), f (full model plus GAD-7*neuroticism_score interaction term). In (g; neuroticism score) and (h; GAD-7) the two strongest predictors of MHQ response were sampled 100 times to test for predicted probability of MHQ participation using the R package effects. In i, we test for multicollinearity among the predictors of MHQ participation using the R package mctest. In j, each intersection of GAD-7 and neuroticism score are characterized.

**Supplementary Table 6.** SNP-based heritability (*h^2^*) estimates from genome-wide association studies of UK Biobank Mental Health Questionnaire (MHQ) responders and non-responders of internalizing outcomes compared to the largest GWAS of a comparable trait in the Million Veteran Program (MVP). Each GWAS also was subjected to multi-trait conditioning and joint analysis using a large GWAS of neuroticism from the Social Science Genetic Association Consortium (SSGAC).

**Supplementary Table 7.** Genetic correlation (*r_g_*) and standard error (SE) between each internalizing trait from the UK Biobank Mental Health Questionnaire (MHQ) responders and non-responders and the largest comparable trait from the Million Veteran Program. Neuroticism and subjective well-being were included as positive and negative *r_g_* controls, respectively. Estimates in grey are the results for a trait pair after subjecting each trait to multi-trait conditioning and joint analysis with neuroticism.

**Supplementary Table 8.** Results of polygenic risk scoring using the --sum-sum method of PRSice v1.25.^62^ GAD and PTSD outcomes ascertained in the UK Biobank Mental Health Questionnaire (MHQ) responders and MHQ non-responders were predicted using a GWAS of a comparable trait from the Million Veteran Program.^5,35^ The UKB MHQ responders and MHQ non-responders were compared using a two-sided Z-test with multiple testing correction adjusting for a false discovery rate (FDR) of 5%.

**Supplementary Table 9.** Comparison of genomic risk loci discovered by GWAS of each internalizing outcome in the UK Biobank Mental Health Questionnaire (MHQ) responders only, MHQ non-responders only, the largest Million Veteran Program GWAS of a comparable trait, meta-analysis of UKB MHQ responders and MHQ non-responders, and meta-analysis of UKB and MVP for each GAD and PTSD outcome. All GWAS were subjected to multi-trait conditioning and joint analysis to account for the effects of neuroticism – as such, MVP risk loci counts are not identical to those reported in their previous publications.^5,35^

**Supplementary Table 10.** Linkage disequilibrium independent genome risk loci discovered by meta-analysis of GAD and posttraumatic stress using two different measures of each trait: a continuous outcome (GAD-7 and PCL-6) and a co-phenome risk score (PheRS). If the genomic risk locus positionally mapped (2-kb window size, 1-kb window on either side^18^) to a genic region, this/these gene names are provided.

**Supplementary Table 11.** Credible set of fine-mapped SNPs associated with each GAD and PTSD outcome and whether the indicated causal SNP is considered the lead SNP in a genomic risk locus.

**Supplementary Table 12.** SNP-based heritability (*h^2^*) of meta-analyzed GAD and PTSD outcomes before and after multi-trait conditioning and joint analysis with neuroticism. Each highlighted GWAS was used as the primary analysis of functional annotation and out-sample polygenic prediction based on the highest conditioned SNP-*h^2^* z-score.

**Supplementary Table 13.** Out-sample polygenic risk scoring using each GAD and PTSD outcome from this study as the base to predict GAD and PTSD outcomes in two target data sets: Yale-Penn and Philadelphia Neurodevelopmental Cohort.

**Supplementary Table 14.** Enrichment and one-sided *P*-values of tissue transcriptomic profiles in various iterations of GAD and PTSD outcome GWAS, including the largest Million Veteran Program studies to date and the meta-analyses from this study (UK Biobank plus MVP). Multiple testing correction was applied using FDR 5% across all traits combined.

**Supplementary Table 15.** Enrichment and one-sided *P*-values of BrainSpan fetal tissue transcriptomic profiles in various iterations of GAD and PTSD outcome GWAS, including the largest Million Veteran Program studies to date and the meta-analyses from this study (UK Biobank plus MVP). Multiple testing correction was applied using FDR 5% across all traits combined.

**Supplementary Table 16.** Functional insights of fetal brain tissue enrichments using Hi-C coupled MAGMA (H-MAGMA) gene-based association tests. Multiple testing correction was applied per GAD and PTSD outcome using a false discovery rate (FDR_Trait_) of 5%. The same multiple testing correction was applied across the entire chromatin interaction analysis for all traits combined (FDR_All_).

**Supplementary Table 17.** All significant cell-type enrichments (one-sided tests) after cross-dataset p-value adjustment. Effect size and standardized effect sizes (BETA_STD) are provided for each cell-type. The cond_state column describes the state of the per dataset conditional analyses based on the cell type from the cond_cell_type column. The step3 column indicates if a cell-type was used for cell-type enrichment analysis step 3 (1) or not (0). Results derived from MAGMA cell-type step 1 and 2.

**Supplementary Table 18.** Results of cell-type cross-data-set conditional analysis showing the relationships between cell-type signals across datasets. Cell types in the same model are conditioned on one another producing cross-dataset beta, standardized effect, standard error, and P-value estimates. Proportional significance (PS) of the cross-data P-value and the cross-dataset marginal (CDM)-P-value on a log_10_ scale such that $PS=\frac{{-log}_{10}\left( P \right)}{{-log}_{10}\left( CDM-P \right)}$. A cell-type pair annotated with “NA” indicates collinearity of signals from these cell-types.

**Supplementary Table 19.** Genetic correlation (*r_g_*) between GAD and PTSD outcomes and medication use phenotypes. Comparison between the *r_g_* estimates using GAD and PTSD outcome GWAS with and without conditioning with neuroticism.

**Supplementary Table 20.** Genetic causality proportions (gĉp) between GAD and PTSD outcomes and medication use phenotypes. Z columns indicate the Z-score for the gĉp such that zscore>>0 implies gĉp>0; P indicates the two-sided p-value for the null hypothesis that gĉp =0; gĉp is the posterior mean gĉp estimate such that gĉp=1 indicates that trait 1 is fully causal for trait 2 and gĉp=-1 indicates the opposite; gcp.SE is the standard error of the gĉp estimate; rho and rho.SE are the genetic correlation and standard error between trait 1 and 2; two p.fully.causal columns provide the p-values for the null hypotheses that gĉp=1 (p.fully.causal.1) or gĉp=-1 (p.fully.causal.2); two *h^2^* columns provide the *h^2^* z-scores for trait 1 (h2.1) and trait 2 (h2.2) – LCV developers recommend interpreting gĉp estimates only when both traits have *h^2^* z-scores >7 (a stringent threshold).

**Supplementary Table 21.** Summary of Mendelian randomization between GAD-2 and vasodilator use using Million Veteran Program GAD-2 data and UKB vasodilator use data. Four analyses are shown testing the bidirectional causal relationship using genome-wide significant SNPs only (GAD-2 🡪 Vasodilator Use and Vasodilator Use 🡪 GAD-2) and all linkage disequilibrium independent SNPs (GAD-2 🡪 Vasodilator Use and Vasodilator Use 🡪 GAD-2). Each causal inference was performed using the robust adjusted profile score to account for weak genetic instruments included in the analysis (i.e., those variants with GWAS P-values > 5x10^-8^) and contains statistics for tests of heterogeneity and horizontal pleiotropy among the genetic instruments.

**Supplementary Table 22.** Genes selected for drug targeting and enrichment analyses were based on having a minGWASp < 5x10^-8^, retention of the highest CADD score in a genomic risk locus, and retention of the gene with the highest probability of loss of function intolerance (pLI score).

**Supplementary Table 23.** Gene Ontology enrichment. For each GAD and PTSD outcome, false discovery rate (5%) adjusted P-values are provided for each functional category.

**Supplementary Table 24.** Results of drug repurposing analyses from the Gene2drug computational tool.^60^ Multiple testing correction was performed using a false discovery rate of 5% per GAD and PTSD outcome.
